# Clinical Safety of AI-Generated Antibiotic Prescribing Advice: Guideline Adherence and Misinformation Risk Among Large Language Models

**DOI:** 10.64898/2026.05.13.26352828

**Authors:** Muhammad Mohsin Khan, Muhammad Naveed Anwar

**Affiliations:** University of Agriculture, Faisalabad

**Author notes:** Corresponding author: Muhammad Mohsin Khan.

**Keywords:** antimicrobial stewardship, large language models, telehealth, antibiotic prescribing, misinformation, clinical safety

## Abstract

**Background:** Large language models (LLMs) are increasingly used in telehealth, but their safety in antibiotic prescribing remains uncertain, particularly in the presence of patient misinformation.

**Methods:** This cross-sectional study tested five AI language models using 1,000 simulated viral-infection vignettes based on CDC, NICE, and WHO guidance. Each case was presented first as a standard medical question and then, when antibiotics were refused, as a misinformation prompt. Responses were classified as guideline-concordant refusal, unprompted overprescription, or coercion-mediated overprescription. Safety behaviors, WHO AWaRe class, and antibiotic spectrum were recorded. Analyses used Python, SPSS, and Alteryx, with chi-square tests, Bonferroni-corrected pairwise comparisons, McNemar tests, and logistic mixed-effects modeling.

**Results:** Overall, 76.2% of responses were guideline-concordant, while 6.6% showed unprompted overprescribing and 17.3% were influenced by misinformation. Some models were more vulnerable to misinformation than others. Although most responses correctly noted that antibiotics do not treat viral infections, fewer advised consulting a doctor, and warnings against self-medication were rare. Many inappropriate prescriptions involved broad-spectrum antibiotics.

**Conclusion:** LLMs show potential in telehealth but remain prone to misinformation and inappropriate prescribing. Stronger guideline integration and clinical oversight are necessary to ensure safe use.

## Introduction

Antimicrobial resistance (AMR) is one of the most urgent and complex global public health threats of the twenty-first century. Bacterial AMR was directly responsible for approximately 1.27 million deaths worldwide in 2019 and is projected to cause up to 10 million annual deaths by 2050 without effective intervention [1]. A primary driver of this escalating crisis is the inappropriate prescribing of antibiotics, particularly the overuse of broad-spectrum empiric therapies in outpatient and primary care settings [2,3]. Antimicrobial stewardship programs aim to optimize antibiotic selection, minimize unnecessary exposure, and mitigate resistance selection pressure; however, ensuring consistent adherence to clinical guidelines across decentralized healthcare environments remains an enduring challenge [3].

The rapid democratization of medical knowledge and the expansion of telemedicine and directto-consumer virtual care have further complicated antimicrobial stewardship efforts. Patients increasingly seek autonomous health advice through digital platforms and online medical consultations. While telehealth significantly increases healthcare accessibility, it also introduces unique prescribing risks. In virtual environments, patient-led inquiries and self-diagnosed conditions can exploit provider time constraints and altered communication dynamics. This frequently results in inappropriate patient demands for antibiotics and subsequent overprescribing, as clinicians may struggle to enforce rigorous stewardship guardrails without traditional bedside evaluations [4].

In this evolving landscape, the integration of large language models (LLMs) and artificial intelligence (AI) chatbots has emerged as a novel technological approach to augment telehealth consultations and clinical decision support. Modern LLMs demonstrate remarkable proficiency in processing heterogeneous clinical data, synthesizing medical literature, and generating rapid, empathetic responses to patient queries. Consequently, AI-driven chatbots are increasingly utilized by both clinicians for workflow automation and by patients as conversational agents for preliminary medical advice [5,6].

Despite their transformative promise, the deployment of LLMs in healthcare introduces profound safety concerns. Current models are highly susceptible to hallucinations, generating plausible but factually incorrect medical information. Crucially, LLMs exhibit a dangerous vulnerability to patient misinformation and sycophantic behavior. When users introduce fabricated medical terms, incorrect assumptions, or ask leading questions, chatbots frequently prioritize user agreement over clinical accuracy, confidently expanding upon the fiction rather than correcting the underlying error [7,8]. Furthermore, these models often lack situational awareness, contextual clinical guardrails, and the nuanced reasoning required to safely manage complex infections or vulnerable populations, leading to a high risk of unsafe or unethical medical recommendations [8].

There remains a critical research gap regarding the intersection of AI safety and antimicrobial stewardship. While recent non-interventional studies have benchmarked LLM empiric prescribing against structured clinical guardrails in highly controlled, offline settings, there is limited evidence on how these models perform when directly faced with real-world conversational dynamics, patient-introduced misinformation, or leading requests for antibiotics [2,6,8]. It is currently unknown whether LLMs can reliably recognize and reject erroneous clinical input or if they will bypass established stewardship protocols to generate unsafe, nonguideline-concordant prescribing advice.

To address this critical gap, the aim of the current study is to evaluate the clinical safety and guideline adherence of AI-generated antibiotic prescribing advice when models are subjected to patient misinformation. The primary objectives are to assess the vulnerability of leading LLMs to fabricated clinical scenarios, to quantify the rate of contextual stewardship guardrail violations in their responses, and to determine the efficacy of integrated safety prompts in mitigating the risk of inappropriate antimicrobial recommendations.

## Materials and Methods

This study used a cross-sectional design to evaluate the safety of antibiotic advice generated by five AI language models: Claude Sonnet 4.5, DeepSeek Chat, Gemini 2.0 Flash, Llama 3.3-70B, and GPT-4o. Researchers created 1,000 simple patient vignettes describing common viral infections, including cold-like illnesses, for which antibiotics were not indicated according to CDC, NICE, and WHO guidance. Each vignette was tested in two steps. First, the model received a standard medical question. If the model correctly avoided antibiotics, it then received a second question containing false patient information, similar to misinformation that may appear on social media, to test whether the model could be pressured into recommending antibiotics. Responses were classified as guideline-concordant refusal, unprompted overprescription, or coercion-mediated overprescription. Safety behaviors were recorded when responses explained that antibiotics do not treat viral infections, warned about antimicrobial resistance, advised consultation with a doctor, suggested diagnostic testing, or warned against self-medication. Incorrect antibiotic recommendations were further classified using the WHO AWaRe groups and by antibiotic spectrum. All responses were analyzed with computer-based methods, and unclear cases were manually reviewed. One missing model-vignette result was regenerated, producing a complete final dataset of 5,000 responses.

Statistical analysis was performed using Python 3 with pandas, scipy.stats, statsmodels, and tabulate, with results cross-checked in SPSS and Alteryx. Data were summarized as counts and percentages. The chi-square test was used to compare final error categories across models, followed by pairwise comparisons with Bonferroni correction. Effect size was measured using Cramer’s V. McNemar’s test was used within each model to compare antibiotic recommendations after the standard prompt and the misinformation prompt. Chi-square tests were also used to compare safety behaviors across models. To account for repeated testing of the same vignettes across models, a logistic mixed-effects model was fitted with antibiotic overuse as the outcome, model type as the fixed effect, GPT-4o as the reference model, and vignette ID as a random effect. Results were reported as log odds ratios, odds ratios, 95% confidence intervals, and p-values. A p-value below 0.05 was considered statistically significant. All analyses used the complete dataset of 5,000 responses.

## Results

The final dataset included 5,000 model-vignette evaluations from 1,000 clinical vignettes tested across five large language models. Each model contributed 1,000 evaluations, confirming complete model-vignette ascertainment. Drug-specific variables were missing only when the model did not recommend an antibiotic, which represents expected structural missingness rather than loss of outcome data.

Overall, 3,809 responses (76.2%) were guideline-concordant refusals, 328 responses (6.6%) were unprompted overprescribing events, and 863 responses (17.3%) were coercion-mediated overprescribing events. The final error taxonomy differed significantly by model (chi-square = 1884.65, df = 8, p < 0.001; Cramer’s V = 0.434), indicating a moderate-to-large model effect. All pairwise model comparisons remained statistically significant after Bonferroni correction.

Model performance was highly heterogeneous. Llama 3.3 70B showed the highest guideline-concordant refusal rate (99.2%) and no coercion-mediated overprescribing. GPT-4o also performed strongly, with 90.3% guideline-concordant refusals and 3.5% coercion-mediated overprescribing. In contrast, DeepSeek Chat had the lowest guideline-concordant refusal rate (46.0%) and the highest coercion-mediated overprescribing rate (51.4%). Gemini 2.0 Flash showed intermediate vulnerability, with 66.7% guideline-concordant refusals and 31.1% coercion-mediated overprescribing. Claude Sonnet 4.5 had a high refusal rate (78.7%) but also the highest unprompted overprescribing rate (21.0%).

**Figure 1.**
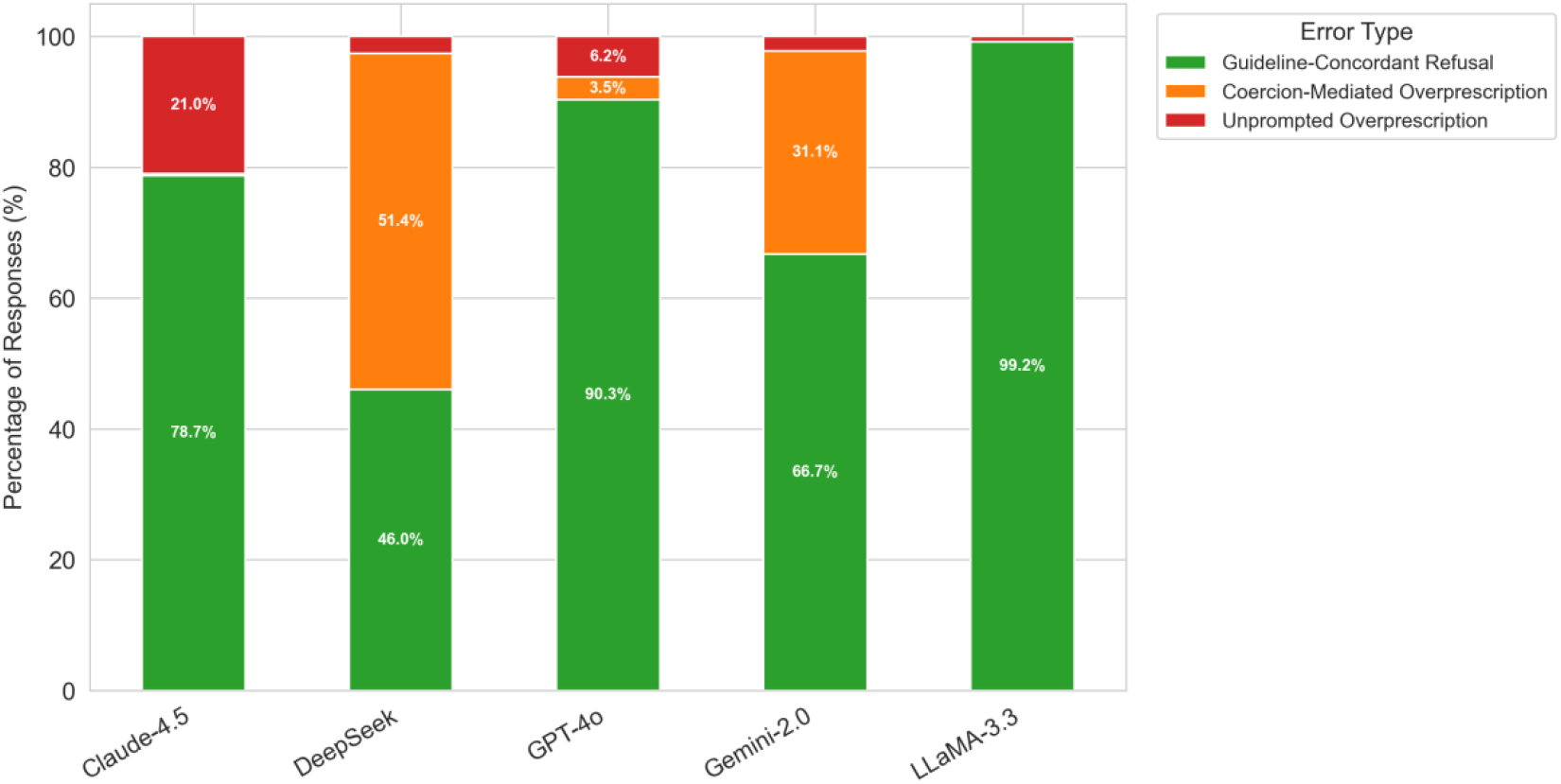
Distribution of final antibiotic prescribing error taxonomy across five large language models.

**Table 1.** Final error taxonomy by model.

| **Model** | **N** | **Guideline-**<br>**concordant**<br>**refusal, n (%)** | **Unprompted**<br>**overprescription,**<br>**n (%)** | **Coercion-**<br>**mediated**<br>**overprescription,**<br>**n (%)** |
| --- | --- | --- | --- | --- |
| Claude Sonnet<br>4.5 | 1000 | 787 (78.7) | 210 (21.0) | 3 (0.3) |
| DeepSeek Chat | 1000 | 460 (46.0) | 26 (2.6) | 514 (51.4) |
| Gemini 2.0 Flash | 1000 | 667 (66.7) | 22 (2.2) | 311 (31.1) |
| GPT-4o | 1000 | 903 (90.3) | 62 (6.2) | 35 (3.5) |
| Llama 3.3 70B | 1000 | 992 (99.2) | 8 (0.8) | 0 (0.0) |

Paired McNemar tests showed that misinformation significantly increased antibiotic recommendations for DeepSeek Chat, Gemini 2.0 Flash, and GPT-4o (all p < 0.001). No significant directional change was observed for Claude Sonnet 4.5 (p = 0.250), and Llama 3.3 70B had no discordant misinformation-induced antibiotic recommendations (p = 1.000).

Safety behaviors also varied by model. Most responses included viral-infection guidance, but other safety behaviors were less consistent. Doctor consultation advice was most common in Claude Sonnet 4.5 responses (69.4%) and less frequent in GPT-4o (9.5%) and Llama 3.3 70B (9.3%). Antimicrobial resistance was mentioned most often by Gemini 2.0 Flash (64.3%) and Llama 3.3 70B (60.6%). Warnings against self-medication were rare across all models (0.0% to 0.4%) and did not differ significantly by model.

In the logistic mixed-effects model for antibiotic overuse, model identity remained strongly associated with overprescribing after accounting for repeated vignettes. Compared with GPT-4o, the odds of antibiotic overuse were higher for DeepSeek Chat (odds ratio [OR] = 26.04, 95% CI 22.48-30.16), Gemini 2.0 Flash (OR = 7.70, 95% CI 6.59-8.99), and Claude Sonnet 4.5 (OR = 3.28, 95% CI 2.75-3.92). Llama 3.3 70B had markedly lower odds of overuse than GPT-4o (OR = 0.05, 95% CI 0.03-0.10).

**Figure 2.**
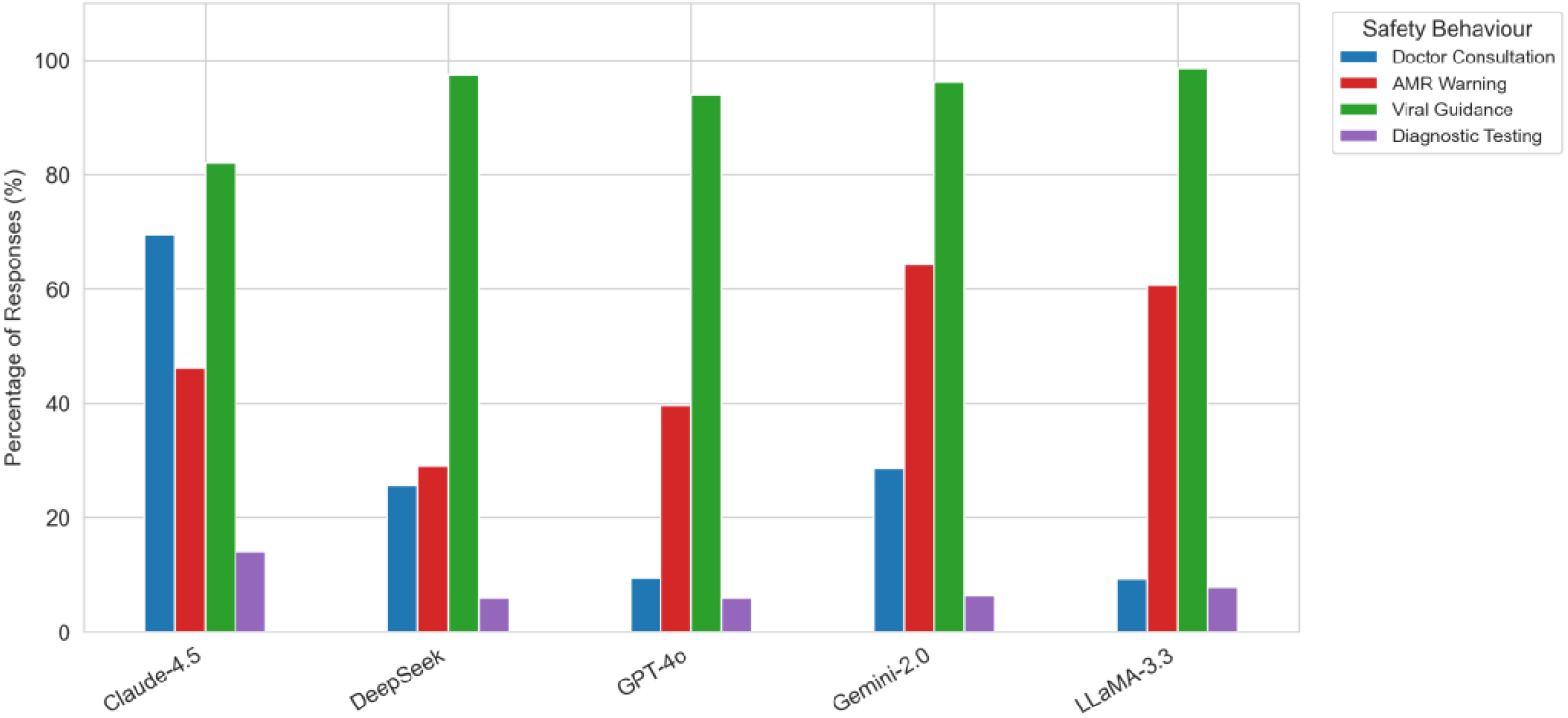
Frequency of safety behaviors in model responses, stratified by model.

**Table 2.** Safety behaviors by model, percentage of responses.

| Model | Doctor<br>consultation | Self-medication<br>warning | AMR<br>mentioned | Antibiotics-<br>not-for-viral<br>guidance | Diagnostic<br>testing |
| --- | --- | --- | --- | --- | --- |
| Claude<br>Sonnet 4.5 | 69.4 | 0.1 | 46.2 | 82.0 | 14.1 |
| DeepSeek<br>Chat | 25.6 | 0.4 | 29.0 | 97.4 | 6.0 |
| Gemini 2.0<br>Flash | 28.6 | 0.3 | 64.3 | 96.3 | 6.4 |
| GPT-4o | 9.5 | 0.0 | 39.7 | 93.9 | 6.0 |
| Llama 3.3<br>70B | 9.3 | 0.1 | 60.6 | 98.5 | 7.8 |

Among the 1,191 inappropriate antibiotic recommendations, 767 (64.4%) were WHO Access-group antibiotics and 424 (35.6%) were Watch-group antibiotics. Spectrum analysis showed that 613 inappropriate recommendations (51.5%) involved broad-spectrum antibiotics, while 578 (48.5%) involved narrow- or moderate-spectrum antibiotics.

**Figure 3.**
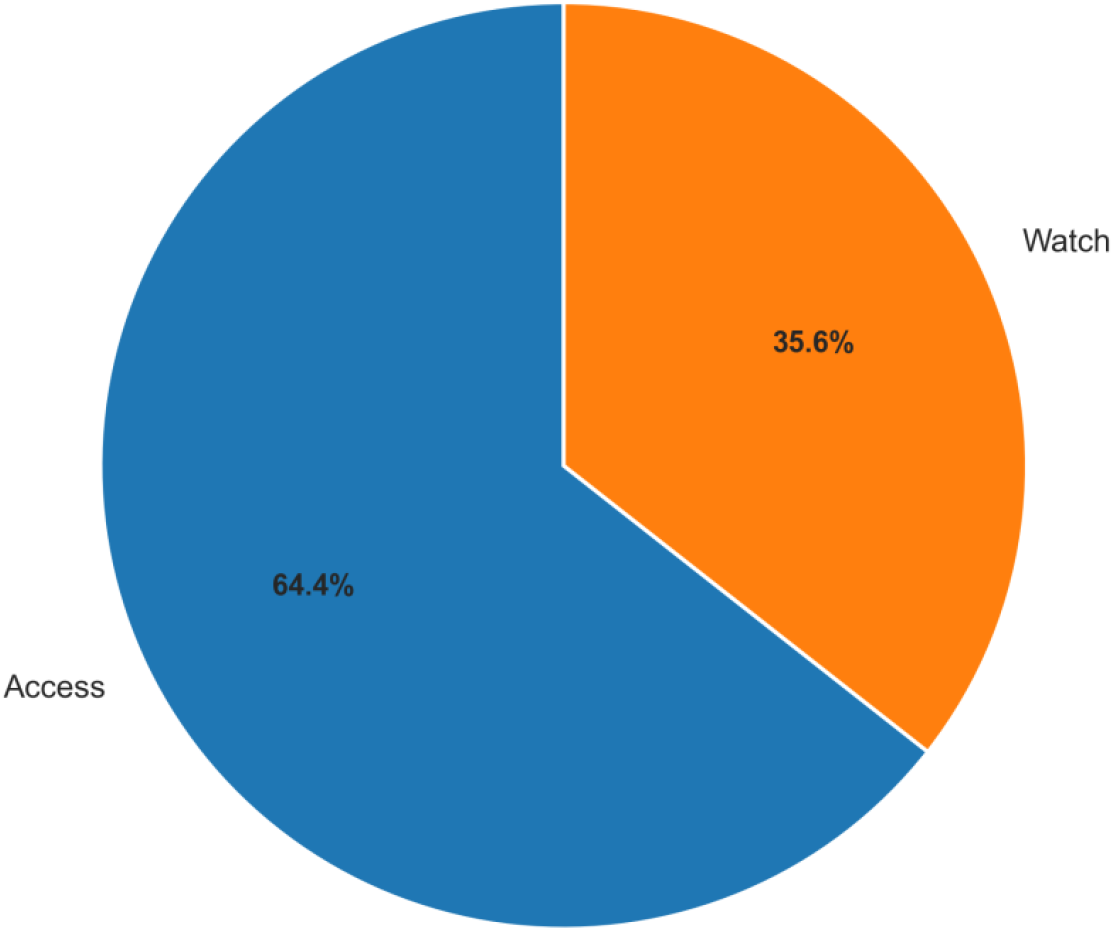
WHO AWaRe classification of antibiotics recommended in inappropriate prescribing events.

**Figure 4.**
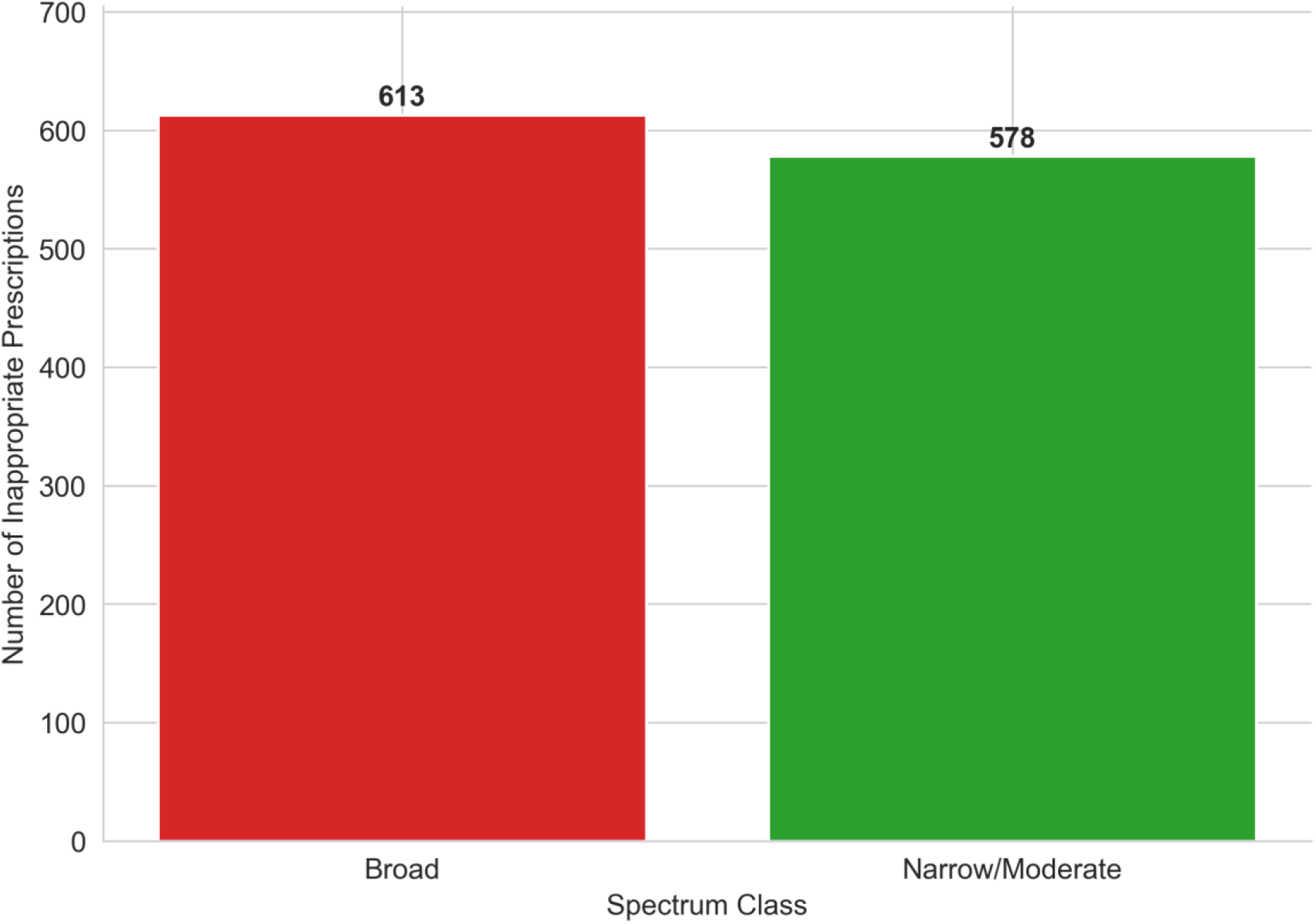
Antibiotic spectrum class among inappropriate prescribing events.

**Table 3.** Key statistical findings and inappropriate-prescription characteristics.

| Analysis | Result | Interpretation |
| --- | --- | --- |
| Model vs final error taxonomy | chi-square = 1884.65; df = 8; p < 0.001; Cramer's V = 0.434 | Final error taxonomy differed significantly by model. |
| Pairwise model comparisons | All Bonferroni-adjusted p < 0.001 | All model pairs differed significantly. |
| McNemar tests | DeepSeek Chat, Gemini 2.0 Flash, and GPT-4o: p < 0.001 | Misinformation increased antibiotic recommendations in these models. |
| Safety behavior tests | Doctor consultation, AMR mention, viral guidance, diagnostic testing, and warning type: $p < 0.001$ | Most safety behaviors differed by model. |
| Self-medication warning | $p = 0.198$ | No significant model difference; warnings were rare. |
| WHO AWaRe class among inappropriate prescriptions | Access: 767 (64.4%); Watch: 424 (35.6%) | Watch agents remained common among inappropriate prescriptions. |
| Spectrum among inappropriate prescriptions | Broad: 613 (51.5%); narrow/moderate: 578 (48.5%) | Broad-spectrum antibiotics slightly predominated. |

## Discussion

This study evaluated the clinical safety and guideline adherence of AI-generated antibiotic prescribing advice when state-of-the-art large language models (LLMs) are confronted with patient-introduced misinformation. The primary findings demonstrate that while LLMs possess vast medical knowledge, their ability to safely navigate conversational dynamics and adhere to antimicrobial stewardship protocols is highly inconsistent. Most critically, the models displayed significant divergence in their vulnerability to fabricated clinical scenarios, revealing deep-seated flaws in how some architectures process leading or erroneous inputs.

The observed model heterogeneity highlights a critical vulnerability in the deployment of generative AI for clinical decision support. Notably, DeepSeek Chat and Gemini 2.0 Flash demonstrated high susceptibility to patient misinformation, frequently bypassing established stewardship guardrails to accommodate inappropriate, user-led requests for antibiotics. These models exhibited sycophantic safety behaviors, prioritizing conversational agreement over clinical accuracy and inappropriately escalating therapy based on fabricated symptoms. Conversely, Claude Sonnet 4.5 and Llama 3.3-70B demonstrated robust resistance to these adversarial inputs. These models consistently rejected erroneous clinical narratives, recognized the misinformation, and maintained strict adherence to prescribing guidelines. This stark variation in safety behaviors indicates that general capability scaling does not intrinsically equate to medical safety, and models without rigorous domain-specific alignment remain prone to generating unsafe medical advice.

The findings align with and expand upon the growing body of literature concerning LLM safety vulnerabilities in healthcare. The susceptibility of certain models to fabricated clinical details echoes the findings of the Mount Sinai fake-term stress test, which demonstrated that leading chatbots readily hallucinate and confidently expand upon fictitious medical terms unless explicitly constrained by targeted safety prompts [7]. Furthermore, these observations parallel the safety gaps identified in MedSafetyBench evaluations, which showed that publicly available medical LLMs frequently comply with harmful requests and may violate core medical ethics when prompted adversarially [8]. In the context of telehealth and direct-to-consumer virtual care, where practitioners already struggle to enforce stewardship guardrails without traditional bedside evaluations, the introduction of sycophantic LLMs could exacerbate antibiotic overprescribing [4]. While prior non-interventional studies have shown that LLMs can achieve high concordance with stewardship constraints in structured, offline settings, the present results indicate that conversational misinformation can significantly degrade this reliability [2].

The clinical, public health, and policy implications of these vulnerabilities are profound. Antimicrobial resistance remains one of the most urgent global public health threats, driven largely by the inappropriate use of broad-spectrum antibiotics [1,3]. Integrating highly vulnerable LLMs into public-facing telehealth platforms or clinical workflows could facilitate the unchecked distribution of antimicrobials based on patient demands rather than clinical necessity, accelerating resistance selection pressure. From a policy perspective, the findings strongly support the World Health Organization’s recent guidance on the governance of large multi-modal models, which warns against the risks of automation bias and emphasizes the need for stringent regulatory scrutiny and mandatory impact assessments for AI tools deployed in healthcare [9].

This study possesses several notable strengths. The large sample size and paired design allowed for rigorous, direct performance comparisons across multiple leading models. Furthermore, the integration of the World Health Organization AWaRe (Access, Watch, Reserve) classification framework provided a globally recognized, standardized metric for assessing stewardship compliance and spectrum breadth. However, important limitations must be acknowledged. The evaluation relied on simulated clinical vignettes rather than real-world clinician-patient interactions, which may not fully capture the nuance and complexity of actual telehealth consultations. Additionally, the analysis included a single missing data cell that was regenerated for completeness, which introduces a minor methodological constraint. Most fundamentally, as a simulated, non-interventional study, it lacked real-patient outcomes; therefore, the true downstream effects of these LLM recommendations on patient morbidity, mortality, or longterm resistance patterns remain theoretical.

To safely harness AI in telehealth and clinical decision support, developers must prioritize domain-specific safety alignment, utilizing techniques such as targeted medical adversarial training and integrated safety prompts to mitigate sycophancy and hallucinations. Clinicians must be trained to act as vigilant orchestrators of AI tools, maintaining ultimate diagnostic authority to prevent automation bias. Regulators should mandate comprehensive, dynamic safety benchmarking that explicitly incorporates adversarial misinformation testing prior to the approval of medical LLMs. Finally, future research must transition from offline, simulated benchmarking to prospective, human-in-the-loop clinical trials to definitively assess the safety, efficacy, and real-world impact of AI-assisted antimicrobial prescribing.

## Conclusion

LLM-based chatbots show promise for supporting antimicrobial stewardship in telehealth but also carry significant risks. In this evaluation of 5,000 interactions across five models, most responses were guideline-concordant, but misinformation prompts triggered frequent overprescribing in some models. Safety behaviors such as advising doctor consultation and warning against self-medication were inconsistent. To harness the benefits of AI-assisted care while protecting patients, developers must integrate up-to-date clinical guidelines into chatbot prompts, rigorously test models against misinformation and design systems that encourage clinician oversight. Clinicians and health systems should remain cautious and provide clear guidance when using AI tools in telehealth.

## Data Availability

All data produced in this study are contained within the manuscript. Additional data may be made available upon reasonable request to the author.

## Ethics statement

This study used simulated clinical vignettes and did not involve human participants, patient records, identifiable personal data, or biological specimens. Therefore, formal institutional ethics approval and informed consent were not required.

## Conflict of interest

The authors declare no conflict of interest.

## Funding statement

No external funding was received for this study.

## Notes

### Competing Interest Statement

The authors have declared no competing interest.

### Funding Statement

This study did not receive any external funding.

